# Enhancing the accuracy of register-based metrics: Comparing methods for handling overlapping psychiatric register entries in Finnish healthcare registries

**DOI:** 10.1101/2023.12.07.23299655

**Authors:** Kimmo Suokas, Mai Gutvilig, Sonja Lumme, Sami Pirkola, Christian Hakulinen

## Abstract

**Objectives:** Healthcare registers are invaluable resources for research. Partly overlapping register entries and preliminary diagnoses may introduce bias. We compare various methods to address this issue and provide fully reproducible open-source R scripts.

**Methods:** We used all Finnish healthcare registers 1969-2020, including inpatient, outpatient and primary care. Four distinct models were formulated based on previous reports to identify actual admissions, discharges, and discharge diagnoses. We calculated the annual number of treatment episodes and patients, and the median length of hospital stay (LOS). We compared these metrics to non-processed data. Additionally, we analyzed the lifetime number of individuals with registered mental disorders.

**Results:** Overall, 2 130 468 individuals had a registered medical contact related to mental disorders. After processing, the annual number of inpatient episodes decreased by 5.13– 10.41% and LOS increased by up to 3 days (27.27%) in years 2011–2020. The number of individuals with lifetime diagnoses reduced by more than 1 percent point (pp) in two categories: schizophrenia spectrum (3.69–3.81pp) and organic mental disorders (1.2– 1.27pp).

**Conclusions:** The methods employed in pre-processing register data significantly impact the number of treatment episodes and LOS. Regarding lifetime incidence of mental disorders, schizophrenia spectrum disorders require a particular focus on data pre-processing.

## Introduction

Healthcare registries provide a valuable resource for medical research due to their ability to encompass large study populations with extensive and continuous follow-up.^1–4^ These registries are generally considered to have good data quality for research purposes.^1,2,5^ However, maintaining the quality of the registers is an ongoing process, marked by various technical nuances. Register-based analyses rely on several methodological assumptions that may compromise reproducibility of the results and challenge international comparisons of healthcare systems.^6^

Registers with continuous and mainly automatized data collection, such as the Finnish healthcare registers, contain temporally overlapping entries and preliminary information. Therefore, a single inpatient treatment episode, for example, may be recorded into multiple register entries.^7^ These multiple entries arise when there are intra-hospital transfers or shifts between distinct medical specialties within the same facility potentially resulting in temporal overlap. Moreover, entries are generated for outpatient and emergency visits that transpire at the outset or during the hospitalization and may contain possibly preliminary diagnoses recorded at the time of these appointments. As a result, combining multiple register entries of different treatment modalities and recognizing potential preliminary diagnoses becomes necessary in order to accurately identify the most reliable estimates of actual discharges, discharge diagnoses, and independent outpatient visits that are not part of inpatient

A few research projects, such as the CEPHOS-LINK project^8^ or the REDD project,^9^ have addressed the importance of the procedures for identifying treatment episodes from the partly overlapping healthcare register data. The CEPHOS-LINK project, for example, indicated that as much as 25% of the register entries associated with psychiatric inpatient care are related to transfers that take place during an ongoing hospitalization.^8^ Despite the prevalence of this issue, a standardized consensus on best practices for handling inpatient episodes within the Finnish registers has yet to be established, leading to variations in criteria applied by different psychiatry-related research projects. For example:

- the CEPHOS-LINK project required that a hospital stay should start and end on distinct calendar days, essentially requiring an inpatient episode to span overnight, whereas others do not have this criterion.^8^ Similarly,
- the REDD project introduced a condition according to which a new treatment period could only commence after a full calendar day spent outside the hospital.^9^ Any entries within the register prior to this transition were amalgamated. This approach aimed to create a clearer distinction between inter-hospital transfers and subsequent rehospitalizations.

To the best of our knowledge, these approaches have not been systematically compared, and the methods employed for pre-processing healthcare registers have generally not been publicly disclosed. This study aims to quantify differences in pre-processing strategies by comparing the number of individuals treated, the number of inpatient episodes, and the average length of stay estimates using different criteria for identifying treatment episodes and discharges. Our analysis incorporates inpatient, secondary outpatient, and primary care data. We hypothesized that different rules for identifying treatment episodes lead to changes in annual and lifetime metrics of psychiatric care with differences across diagnostic categories. Additionally, we provide fully reproducible, open-source R scripts along with example synthetic data, in order to enable others to evaluate and benefit from this effort.

## Method

For this study, all Finnish healthcare register data until the end of 2020 was utilized. Individuals with a history of mental health-related contact with psychiatric inpatient care were reliably recognized since 1975, secondary outpatient care was included since 1998 and primary care since 2011.

The Research Ethics Committee of the Finnish Institute for Health and Welfare approved the study protocol (decision #10/2016§751). Informed consent is not required for register-based studies in Finland.

### Description of the registers

The Finnish Care Register for Health Care (FCRHC), formerly known as the Finnish Hospital Discharge Register (FHDR) prior to 1994, provides continuous nationwide inpatient data with coverage dating back to 1969, making it the first register among Nordic countries.^2,5^ The format of the data is slightly different across the years. Initially, the FHDR consisted of separate sub-registers catering to diverse hospital types, including general, tuberculosis, psychiatric, and others. These sub-registers were subsequently consolidated into a unified system.^5^ For the years 1969-1974, the classification of medical specialties in some sub-registries may lack clarity.

Data on specialized outpatient care in the public sector have been collected since 1998, with consistent comparability across time and service providers achieved from 2006 onward.^10^ The Register of Primary Health Care visits (RPHC) encompasses publicly organized outpatient primary health care since 2011. Its inclusion has significantly enhanced the comprehensiveness of the registers, potentially influencing results in register-based psychiatric epidemiological research, especially due to the common utilization of primary care for mental health treatment in Finland.^11,12^

The International Statistical Classification of Diseases and Related Health Problems, Tenth Revision (ICD-10) has been used in Finland since 1996. Prior to that, the Finnish version of the ICD-9 was used from 1987 to 1995, and ICD-8 from 1969 to 1986. In some primary care facilities, the International Classification of Primary Care, Second Edition (ICPC-2), is used instead of ICD-10.

### Procedures

#### Sample definitions and initial data transformations

To ensure data quality and consistency, the following initial quality controls and data transformations were implemented: entries without a personal identification number were excluded. Entries where the admission or discharge date was missing, discharge was before admission, or entries out of the time range of the dataset were excluded.

Starting from 1996, patients in inpatient care on the last day of the year were reported in the register. In these entries, the discharge day should be left blank; however, this was not consistently observed and was accounted for in the scripts.

The coding of psychiatric care varies across years. Uniform specialty coding in all register entries commenced in 1987, with further coding changes introduced in 1994. Prior to 1987, register entries from both mental hospitals and general hospitals with psychiatry as a specialty were considered psychiatric care.

Prior to 1998, all entries pertained to inpatient care. Subsequently, distinguishing entries related to inpatient services became necessary. The coding of the treatment type underwent substantial changes starting in 2019. In 2019, both the old and new coding systems were in use, and some entries even featured a mixture of both, which needs to be accounted for. In addition, outpatient episodes spanning over more than two calendar days were considered inpatient care. For outpatient and emergency visits, consecutive start and end dates were allowed.

RPHC contains data on assessment of the need for care, scheduling of the appointments, consultations between professionals and beyond. For the purpose of this study, in-person and virtual real-time contacts were included.

Mental health-related diagnoses under ICD-8, ICD-9, and ICPC-2 were converted to corresponding ICD-10 sub-chapter categories whenever possible. Conversion tables provided by the classification developers were utilized when possible.^13,14^ ICPC-2 concepts that did not have exact counterparts in ICD-10 were grouped separately and were not included in the ICD-10 sub-chapter categories.

#### Identification of inpatient episodes and outpatient appointments during the episode

Using the criteria outlined in the introduction section, we have derived four distinct models for identifying treatment episodes from the healthcare registers (Table 1). Inpatient episodes were identified as follows:

**Table 1:** Possible models for identifying inpatient episodes.

| Model | Description |
| --- | --- |
| 1 | A new hospitalization may begin the day after a previous hospitalization ended, with no specific minimum length required for a hospitalization. This represents the most liberal approach |
| 2 | A new hospitalization may begin the day after a previous hospitalization ended. Valid hospitalizations are those that extend over a minimum of two consecutive days, incorporating at least one overnight stay. If both admission and discharge take place on the same day, the visit is classified as an outpatient visit. This model was used in the CEPHOS-LINK project<sup>8</sup> |
| 3 | A new hospitalization is allowed after a full day has been spent outside the hospital after the previous hospitalization ended. There is no specific minimum duration required for a hospitalization. This model was used in the REDD project<sup>9</sup> |
| 4 | A new hospitalization is allowed after a full day has been spent outside the hospital after the previous hospitalization ended. Valid hospitalizations are those that extend over a minimum of two consecutive days, incorporating at least one overnight stay. If both admission and discharge take place on the same day, the visit is classified as an outpatient visit. This represents the most conservative model |

1. We identified overlapping register entries related to inpatient care, with transfers recognized in two different ways: First, discharges and new admissions on the same day were considered transfers during an inpatient episode (Models 1 and 2, Table 1), and second, discharges and new admissions without a full calendar day outside the hospital were considered transfers (Models 3 and 4, Table 1). After that,
2. Overnight stay was examined and for Models 2 and 4, episodes starting and ending on the same day were reclassified as outpatient events.

After identifying inpatient episodes with each model, possible secondary outpatient and primary care appointments during the inpatient episode were identified. These appointments were not considered independent appointments. Hence, the model selected for identifying inpatient episodes affected the number of outpatient and primary care appointments in the fully processed data.

#### Identification of discharge diagnoses and outpatient diagnoses

We recorded discharge diagnoses on the last day of the episode, as well as on the last day of psychiatric inpatient care, if the latter occurred before the final discharge. Additionally, we documented all diagnoses made during the inpatient stay. Outpatient and primary care visits during the course of inpatient episodes were also identified. Psychiatric outpatient diagnoses registered on the day of or the days following discharge from a psychiatric ward were included in the final discharge diagnoses.

Inpatient, outpatient, and primary care diagnoses established before the end of the episode were categorized as preliminary and were not considered discharge diagnoses. If an inpatient episode included psychiatric care, mental disorder diagnoses from specialties other than psychiatry were excluded.

The earliest age at which a person might possibly develop a specific disorder were set in a similar way as previously.^15^

### Analysis

After identifying inpatient episodes and outpatient and primary care events associated with inpatient care, we compared the number of episodes and treated individuals calculated with the four models to corresponding numbers from the non-processed data (with initial data transformations only).

First, we conducted separate analyses for the years 2011-2020 to assess the impact of episode identification on annual descriptive statistics, including the number of psychiatric inpatient episodes, median length of stay in psychiatric inpatient care, and the number of treated individuals within specific diagnostic categories.

Second, we calculated the total number of individuals within specific diagnostic categories between 1975 and 2020.

Numbers and differences in percentages are presented throughout the study. We used R, version 4.2.2 for all data processing. The scripts have been made publicly available and contain supplementary description of each step of the process summarized above (https://github.com/kmmsks/hilmo_identify_episodes/).

## Results

Between the years 1975 and 2020, 2 130 468 individuals had a valid registered medical contact related to mental disorders. Between years 2011 and 2020, less than 0.4% of observations were excluded due to missing personal identification number, missing admission date or discharge date, discharge recorded before admission, or entries out of the time range of the data set.

### Annual number of psychiatric episodes and patients

After the identification of inpatient episodes, accounting for the overlapping days, and recognizing outpatient appointments during inpatient care, the most liberal model (Model 1) resulted in 5.13–7.08% less inpatient episodes than non-processed data. Applying the most conservative model (Model 4) for inpatient care reduced the numbers of episodes by additional 2.33–3.72pp (Table 2).

**Table 2:**
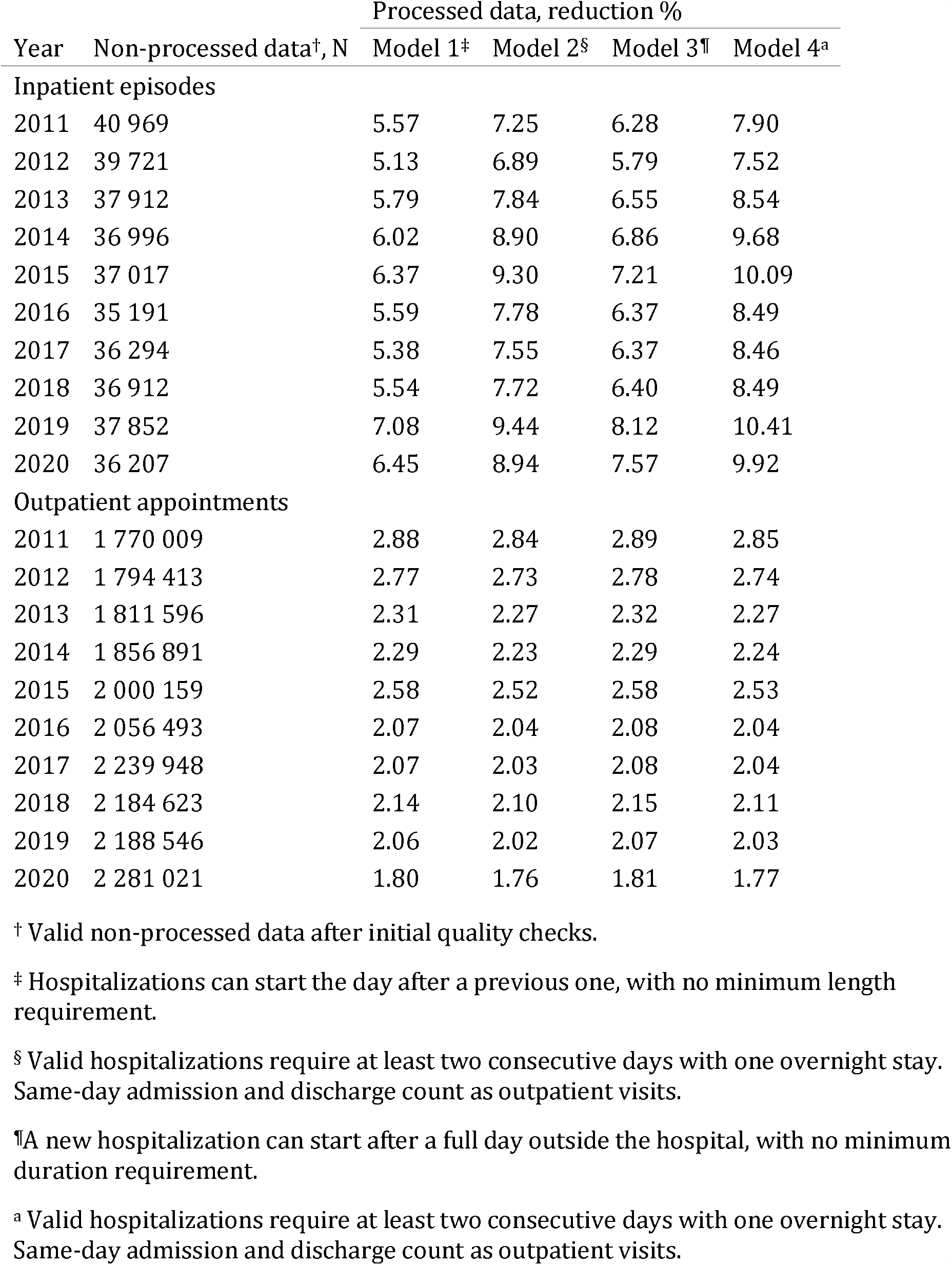
Annual number of register entries related to psychiatric care and reduction in the number of inpatient and outpatient episodes after identification of episodes with different models.

Outpatient episodes were recognized after the identification of inpatient episodes. Model 1 resulted in 1.76–2.84% less outpatient events compared to non-processed data. There were practically no difference between the models (less than 0.1pp) (Table 2).

The annual number of individuals with inpatient treatments in the year 2020 is shown in Table 3. If an overnight stay was required for a visit to count as inpatient treatment, the number of individuals with inpatient care decreased by 1.51–1.50%. Diagnosis-specific values changed by less than 3% in all ICD-10 sub-chapter categories. It is worth noticing that the number of individuals increased with pre-processing when psychiatric outpatient visits during an inpatient stay were included.

**Table 3:**
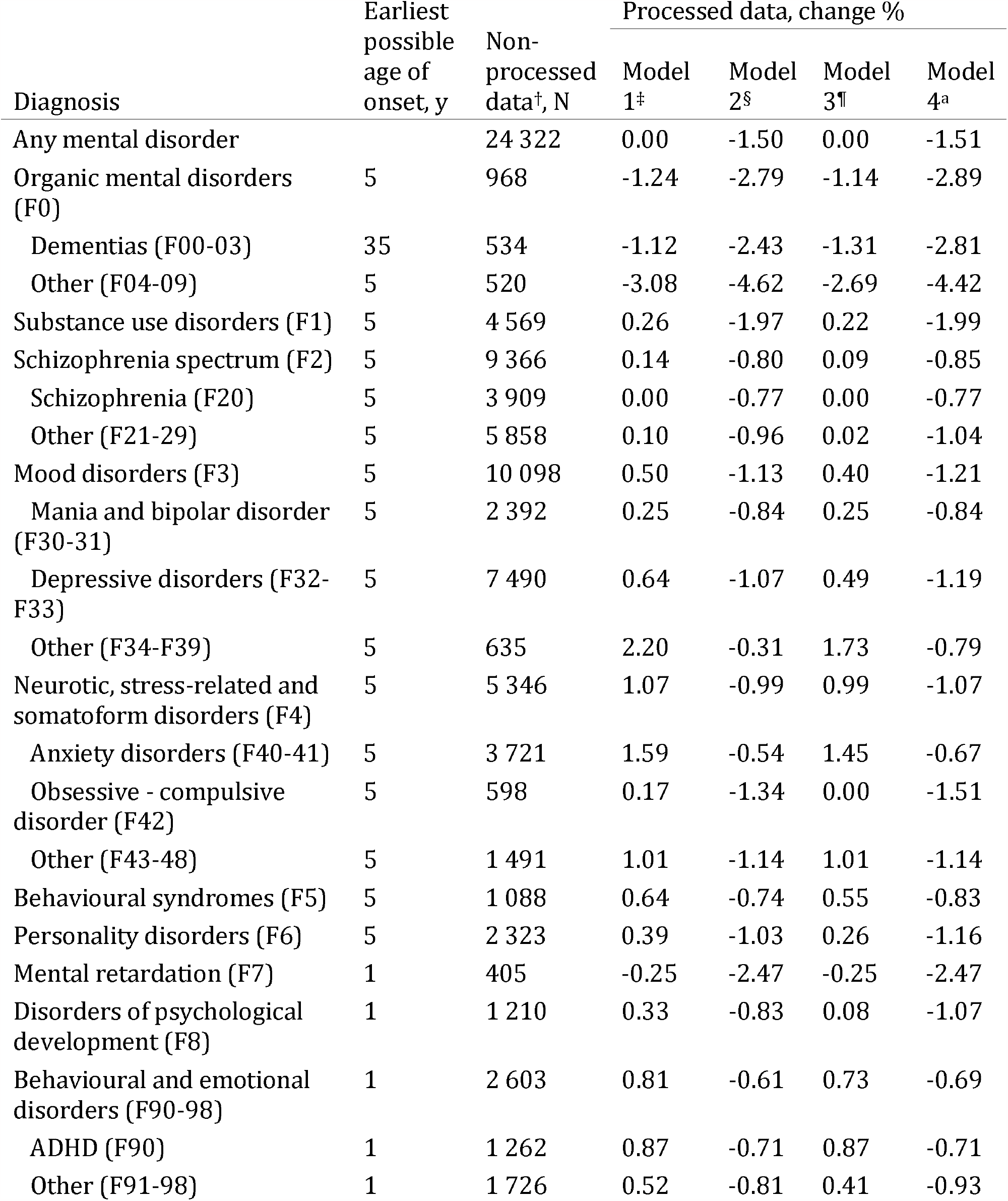
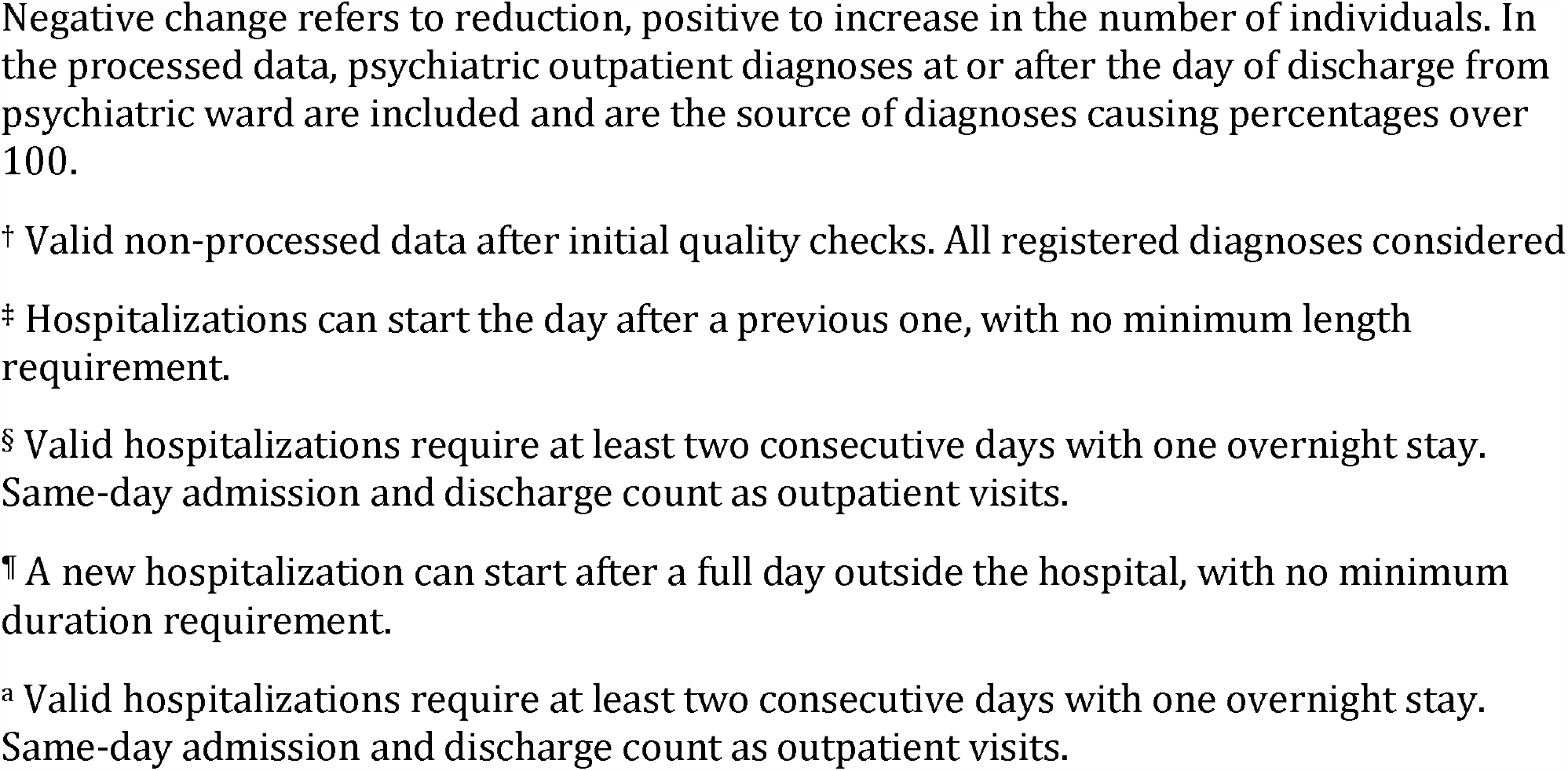
Annual number of individuals with psychiatric inpatient care by discharge diagnosis in non-processed data and after identification of inpatient episodes with different models in 2020.

| Diagnosis | Earliest possible age of onset, y | Non-processed data<sup>†</sup>, N | Processed data, change % |  |  |  |
| --- | --- | --- | --- | --- | --- | --- |
|  |  |  | Model 1<sup>‡</sup> | Model 2<sup>§</sup> | Model 3<sup>¶</sup> | Model 4<sup>a</sup> |
| Any mental disorder |  | 24 322 | 0.00 | -1.50 | 0.00 | -1.51 |
| Organic mental disorders (F0) | 5 | 968 | -1.24 | -2.79 | -1.14 | -2.89 |
| Dementias (F00-03) | 35 | 534 | -1.12 | -2.43 | -1.31 | -2.81 |
| Other (F04-09) | 5 | 520 | -3.08 | -4.62 | -2.69 | -4.42 |
| Substance use disorders (F1) | 5 | 4 569 | 0.26 | -1.97 | 0.22 | -1.99 |
| Schizophrenia spectrum (F2) | 5 | 9 366 | 0.14 | -0.80 | 0.09 | -0.85 |
| Schizophrenia (F20) | 5 | 3 909 | 0.00 | -0.77 | 0.00 | -0.77 |
| Other (F21-29) | 5 | 5 858 | 0.10 | -0.96 | 0.02 | -1.04 |
| Mood disorders (F3) | 5 | 10 098 | 0.50 | -1.13 | 0.40 | -1.21 |
| Mania and bipolar disorder (F30-31) | 5 | 2 392 | 0.25 | -0.84 | 0.25 | -0.84 |
| Depressive disorders (F32-F33) | 5 | 7 490 | 0.64 | -1.07 | 0.49 | -1.19 |
| Other (F34-F39) | 5 | 635 | 2.20 | -0.31 | 1.73 | -0.79 |
| Neurotic, stress-related and somatoform disorders (F4) | 5 | 5 346 | 1.07 | -0.99 | 0.99 | -1.07 |
| Anxiety disorders (F40-41) | 5 | 3 721 | 1.59 | -0.54 | 1.45 | -0.67 |
| Obsessive - compulsive disorder (F42) | 5 | 598 | 0.17 | -1.34 | 0.00 | -1.51 |
| Other (F43-48) | 5 | 1 491 | 1.01 | -1.14 | 1.01 | -1.14 |
| Behavioural syndromes (F5) | 5 | 1 088 | 0.64 | -0.74 | 0.55 | -0.83 |
| Personality disorders (F6) | 5 | 2 323 | 0.39 | -1.03 | 0.26 | -1.16 |
| Mental retardation (F7) | 1 | 405 | -0.25 | -2.47 | -0.25 | -2.47 |
| Disorders of psychological development (F8) | 1 | 1 210 | 0.33 | -0.83 | 0.08 | -1.07 |
| Behavioural and emotional disorders (F90-98) | 1 | 2 603 | 0.81 | -0.61 | 0.73 | -0.69 |
| ADHD (F90) | 1 | 1 262 | 0.87 | -0.71 | 0.87 | -0.71 |
| Other (F91-98) | 1 | 1 726 | 0.52 | -0.81 | 0.41 | -0.93 |

Negative change refers to reduction, positive to increase in the number of individuals. In the processed data, psychiatric outpatient diagnoses at or after the day of discharge from psychiatric ward are included and are the source of diagnoses causing percentages over 100.
† Valid non-processed data after initial quality checks. All registered diagnoses considered
¶ A new hospitalization can start after a full day outside the hospital, with no minimum duration requirement.

### Length of stay in psychiatric inpatient care

The median length of stay increased by 1 to 3 days in different years with different models compared to the non-processed data (Table 4). In 2013, the difference was the greatest, 3 days, resulting in a 27.27% change in the median length of stay. In 2020, the median length of stay increased from 8 to 9 days (12.5%) in all models. The distribution of the number of episodes by length of stay is shown in Figure 1.

**Table 4:** Annual median length of stay in psychiatric inpatient care in non-processed data and after identification of inpatient episodes with different models.

| Year | Non-processed data<sup>†</sup> | Processed data |  |  |  |
| --- | --- | --- | --- | --- | --- |
|  |  | Model 1<sup>‡</sup> | Model 2<sup>§</sup> | Model 3<sup>¶</sup> | Model 4<sup>a</sup> |
| 2011 | 12 | 13 | 14 | 13 | 14 |
| 2012 | 12 | 13 | 14 | 13 | 14 |
| 2013 | 11 | 13 | 13 | 13 | 14 |
| 2014 | 11 | 12 | 13 | 13 | 13 |
| 2015 | 11 | 12 | 13 | 12 | 13 |
| 2016 | 11 | 12 | 13 | 12 | 13 |
| 2017 | 11 | 11 | 12 | 11 | 12 |
| 2018 | 10 | 11 | 12 | 11 | 12 |
| 2019 | 9 | 10 | 11 | 10 | 11 |
| 2020 | 8 | 9 | 9 | 9 | 9 |

<sup>†</sup> Valid non-processed data after initial quality checks.
<sup>‡</sup> Hospitalizations can start the day after a previous one, with no minimum length requirement.
<sup>§</sup> Valid hospitalizations require at least two consecutive days with one overnight stay. Same-day admission and discharge count as outpatient visits.
<sup>¶</sup> A new hospitalization can start after a full day outside the hospital, with no minimum duration requirement.
<sup>a</sup> Valid hospitalizations require at least two consecutive days with one overnight stay. Same-day admission and discharge count as outpatient visits.

**Figure 1.**
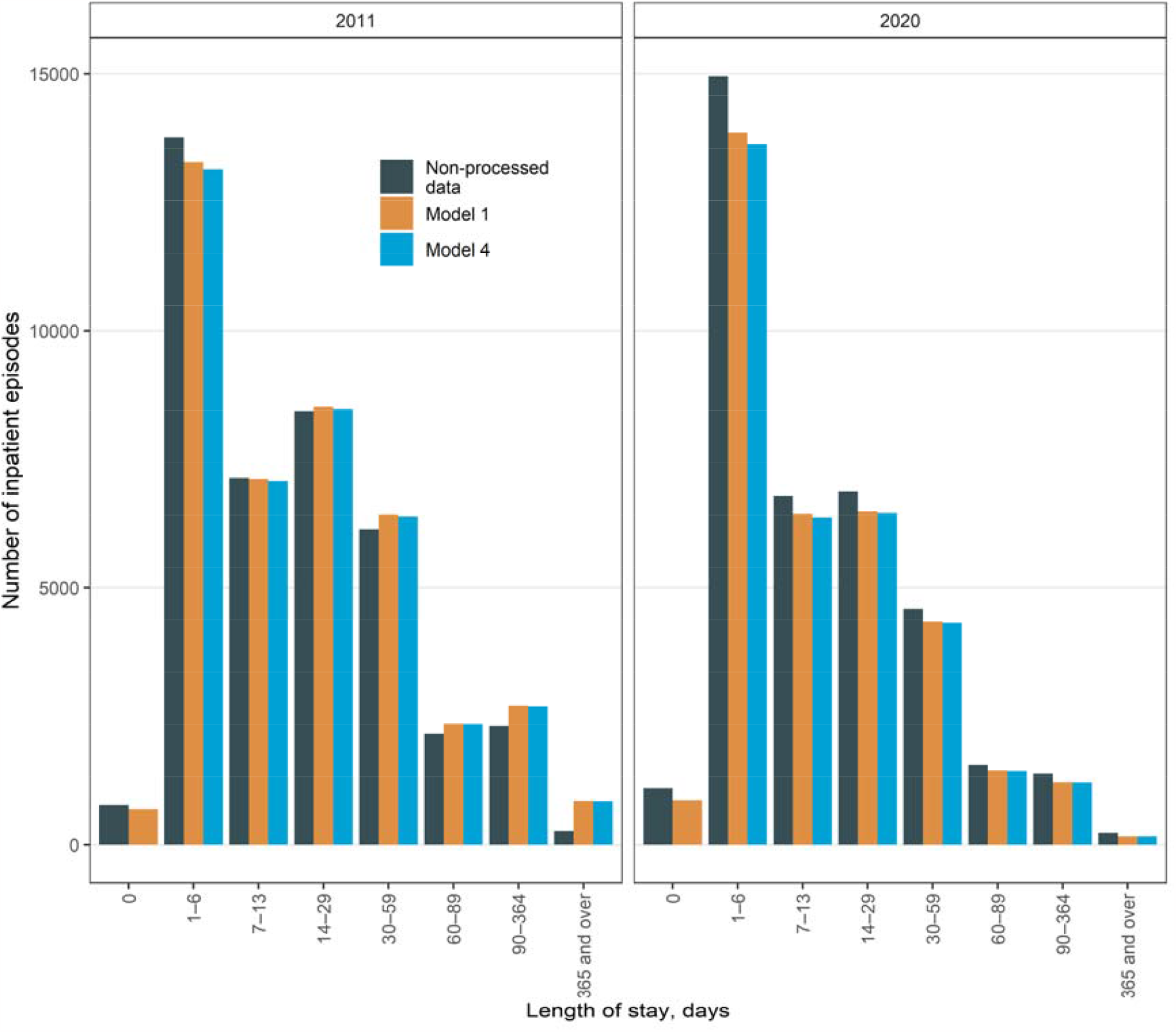
Lenght of stay of psychiatric inpatient care in the years 2011 and 2020. Categorization is based on the official reports. Non-processed data refers to valid non-processed data after initial quality checks. Model 1 refers to hospitalizations can start the day after a previous one, with no minimum length requirement. Model 4 refers to valid hospitalizations require at least two consecutive days with one overnight stay. same-day admission and discharge count as outpatient visits.

### Overall number of individuals covered by diagnosis

Between the years 1975 and 2020, 2 130 468 individuals had medical contacts related to mental disorders (Table 5). After the identification of treatment episodes, the number of individuals with a life-time diagnoses was reduced by more than 1pp only in two of the ICD-10 sub-chapter categories: schizophrenia spectrum disorders (F20-F29) by 3.69– 3.81pp and organic, including symptomatic, mental disorders (F00-09) by 1.2–1.27pp. More specifically, organic mental disorders other than dementias (ICD-10: F04-09) as well as unspecified nonorganic psychosis (F29) and acute and transient psychotic disorders (F23) were noteworthy sources of variation in these sub-chapter categories (Table 5).

**Table 5:** Overall number of individuals with registered mental disorders by diagnosis in non-processed data and after identification of inpatient episodes with different models, years 1975–2020 and 1996–2020.

| Diagnosis | Earliest possible age of onset, y | Non-processed data<sup>†</sup>, N | Processed data, reduction % |  |  |  |
| --- | --- | --- | --- | --- | --- | --- |
|  |  |  | Model 1<sup>‡</sup> | Model 2<sup>§</sup> | Model 3<sup>¶</sup> | Model 4<sup>a</sup> |
| Any mental disorder |  | 2 130 468 | 0.01 | 0.01 | 0.01 | 0.01 |
| Organic mental disorders (F0) | 5 | 197 915 | 1.20 | 1.20 | 1.27 | 1.27 |
| Dementias (F00-03)<sup>b</sup> | 35 | 114 295 | 0.48 | 0.48 | 0.51 | 0.51 |
| Other (F04-09)<sup>b</sup> | 5 | 47 271 | 4.93 | 4.93 | 5.14 | 5.14 |
| Substance use disorders (F1) | 5 | 286 587 | 0.65 | 0.65 | 0.68 | 0.68 |
| Schizophrenia spectrum (F2) | 5 | 186 718 | 3.69 | 3.69 | 3.81 | 3.81 |
| Schizophrenia (F20)<sup>b</sup> | 5 | 53 985 | 1.95 | 1.95 | 2.02 | 2.02 |
| Schizotypal disorder (F21)<sup>b</sup> | 5 | 7 079 | 3.83 | 3.83 | 3.96 | 3.96 |
| Persistent delusional disorders (F22)<sup>b</sup> | 5 | 28 820 | 5.06 | 5.06 | 5.22 | 5.22 |
| Acute and transient psychotic disorders (F23)<sup>b</sup> | 5 | 24 783 | 9.78 | 9.78 | 10.06 | 10.06 |
| Induced delusional disorder (F24)<sup>b</sup> | 5 | 223 | 14.35 | 14.35 | 14.35 | 14.35 |
| Schizoaffective disorders (F25)<sup>b</sup> | 5 | 16 123 | 3.09 | 3.09 | 3.19 | 3.19 |
| Other nonorganic psychotic disorders (F28)<sup>b</sup> | 5 | 3 728 | 5.95 | 5.95 | 6.12 | 6.12 |
| Unspecified nonorganic psychosis (F29)<sup>b</sup> | 5 | 66 166 | 16.34 | 16.34 | 16.71 | 16.71 |
| Mood disorders (F3) | 5 | 654 874 | 0.66 | 0.66 | 0.68 | 0.68 |
| Mania and bipolar disorder (F30-31)<sup>b</sup> | 5 | 64 112 | 1.79 | 1.79 | 1.83 | 1.83 |
| Depressive disorders (F32-F33)<sup>b</sup> | 5 | 585 175 | 0.72 | 0.72 | 0.73 | 0.73 |
| Other (F34-F39)<sup>b</sup> | 5 | 78 838 | 1.59 | 1.59 | 1.64 | 1.64 |
| Neurotic, stress-related and somatoform disorders (F4) | 5 | 760 972 | 0.42 | 0.42 | 0.45 | 0.45 |
| Anxiety disorders (F40-41)<sup>b</sup> | 5 | 404 288 | 0.65 | 0.65 | 0.66 | 0.66 |
| Obsessive - compulsive disorder (F42)<sup>b</sup> | 5 | 28 918 | 0.54 | 0.54 | 0.57 | 0.57 |
| Other (F43-48)<sup>b</sup> | 5 | 361 708 | 0.49 | 0.49 | 0.51 | 0.51 |
| Behavioural syndromes (F5) | 5 | 359 745 | 0.38 | 0.38 | 0.38 | 0.38 |
| Personality disorders (F6) | 5 | 134 291 | 0.79 | 0.79 | 0.86 | 0.86 |
|  |  |  | Model 1<sup>‡</sup> | Model 2<sup>§</sup> | Model 3<sup>¶</sup> | Model 4<sup>a</sup> |
| Mental retardation (F7) | 1 | 27 259 | 0.55 | 0.55 | 0.59 | 0.59 |
| Disorders of psychological development (F8) | 1 | 126 565 | 0.69 | 0.69 | 0.76 | 0.76 |
| Behavioural and emotional disorders (F90-98) | 1 | 269 365 | 0.55 | 0.55 | 0.56 | 0.56 |
| ADHD (F90)<sup>b</sup> | 1 | 79 027 | 0.10 | 0.10 | 0.10 | 0.10 |
| Other (F91-98)<sup>b</sup> | 1 | 114 770 | 0.40 | 0.40 | 0.41 | 0.41 |

<sup>†</sup> Valid non-processed data after initial quality checks.
<sup>‡</sup> Hospitalizations can start the day after a previous one, with no minimum length requirement.
<sup>§</sup> Valid hospitalizations require at least two consecutive days with one overnight stay. Same-day admission and discharge count as outpatient visits.
<sup>¶</sup> A new hospitalization can start after a full day outside the hospital, with no minimum duration requirement.
<sup>a</sup> Valid hospitalizations require at least two consecutive days with one overnight stay. Same-day admission and discharge count as outpatient visits.
<sup>b</sup> Specific diagnoses only for the period of 1996–2020, to avoid inaccuracy in conversion of ICD-8 and ICD-9 diagnoses.

## Discussion

This study demonstrated that the pre-processing of partly overlapping register entries is a complex process that has important implications for the scientific results and administrative metrics of psychiatric care derived from healthcare registers. The annual count of inpatient episodes showed a reduction ranging from 5% to 10%, while the median length of stay increased by up to 27% following the identification of inpatient episodes. However, the overall number of individuals with registered mental disorders in psychiatric services or primary care remained relatively stable, with two notable exceptions: the exclusion of preliminary diagnoses proved to be of clear significance when assessing psychotic disorders or organic mental disorders. To enhance the precision of register-based analyses, the management of overlapping register entries should be carried out and reported systematically. We have introduced a reproducible open-source method for this process.

The methods employed in this study identify days in inpatient care that are covered in multiple register entries with high certainty. Therefore, using any of these models improves accuracy of any metrics derived from the registers. On the other hand, selecting the most conservative model instead of the most liberal, yielded only minor further adjustments in the estimates. While these models are likely to overlook a small proportion of patients with same-day re-hospitalizations, this has been regarded as a lesser concern compared to erroneously categorizing transfers as readmissions.^6^ Whether there is a need for a minimum length requirement for a hospitalization is a subjective judgment, as the registers do not provide a definitive answer, given their inability to discern the reasons for episodes lasting less than a day. The relevance of a minimum length criterion depends on the primary focus of the analysis; for instance, if the emphasis is on admissions, a minimum length might not be advantageous, whereas if inpatient treatments are the primary focus, it could be justified. With the provided scripts, however, these criteria can be easily customized.

The issue of partly overlapping register entries and preliminary diagnoses remains relatively underexplored in register-based research. Some researchers have addressed this by excluding diagnoses from emergency departments and by eliminating duplicate entries.^16,17^ Finnish official statistics report an annual median length of stay in psychiatric inpatient care that closely aligns with our non-processed statistics, with the only exception of 2017, where there is a one-day discrepancy.^18^ However, after the identification of inpatient episodes, our estimates for the length of stay tend to be one to three days higher. The provision of open-source methodologies is instrumental in achieving reproducible and unequivocal analyses and plays a role in promoting open science.^19,20^

Identifying of discharge diagnoses displayed variation across different diagnostic categories, with schizophrenia and organic disorders exhibiting the highest prevalence of preliminary diagnoses. This observation aligns with expectations, as the diagnoses of psychotic disorders are known to frequently evolve during follow-up.^21–23^ A landmark study indicated that although the Finnish healthcare registers are effective in screening for possible psychotic disorders, they may not be optimal for complete case ascertainment.^24^ Therefore, the application of conservative models for the exclusion of preliminary diagnoses may prove advantageous. Conversely, it is worth noting that during the 1990s, a trend toward a more restrictive definition of schizophrenia in clinical practice in Finland was recognized, potentially resulting in a higher likelihood of false negatives and fewer false positives.^25^ However, the accuracy of diagnostic practices in primary or secondary care mental health services has not been evaluated recently in Finland. Diagnostics in psychiatry is a complex process with clinical and administrative considerations.^26^ Accordingly, the unspecified category (F29) emerged as the most prevalent diagnosis among all the disorders within the ICD-10 sub-chapter Schizophrenia, schizotypal and delusional disorders before and after the processing of the data.

### Limitations

First, this study solely relied on register data and lacked clinical or other reference data for performance comparison among the models. While all models successfully identify overlapping days, we lacked the means to determine the superiority of any model in recognizing hospital transfers resulting from early readmissions. Second, individual patients may have intricate inpatient care pathways involving multiple medical specialties. The general procedures outlined in this study may not offer sufficient detail if the focus is on diagnosing the length of stay within specific specialties in such cases. Instead, further development of methods is warranted. Third, present results are based on Finnish psychiatric register data and further studies are needed to replicate the present findings in other settings.

## Conclusions

Register-based research benefits from constant methodological development. Technical intricacies, including those related to partly overlapping register entries, may prove complex and labor-intensive. While the issues tied to partly overlapping register entries may be central to certain research questions, the actual scientific interest may often lay elsewhere. The application of open science principles and collaborative development of readily available solutions can significantly save time and enhance research quality by improving comparability and reproducibility of results.

## Data Availability

The data that support the findings of this study are available from the National Institute of Health and Welfare (www.thl.fi) and Statistics Finland (www.stat.fi). Restrictions apply to the availability of these data, which were used under license for this study. Inquiries about secure access to data should be directed to data permit authority Findata (findata.fi/en). The method described in this article has been made publicly available and contain supplementary description of each step of the process (https://github.com/kmmsks/hilmo_identify_episodes/).

## Acknowledgments

Kimmo Suokas was supported by the Jalmari and Rauha Ahokas Foundation and the Finnish Psychiatric Association. Christian Hakulinen was supported by the Academy of Finland (354237) and the European Union (ERC, MENTALNET, 101040247). Views and opinions expressed are however those of the authors only and do not necessarily reflect those of the European Union or the European Research Council. Neither the European Union nor the granting authority can be held responsible for them.

## Conflict of Interest Statement

We declare no competing interests.

## Notes

**Funding statement:** Kimmo Suokas was supported by the Jalmari and Rauha Ahokas Foundation and the Finnish Psychiatric Association. Christian Hakulinen was supported by the Academy of Finland (354237) and the European Union (ERC, MENTALNET, 101040247). Views and opinions expressed are however those of the authors only and do not necessarily reflect those of the European Union or the European Research Council. Neither the European Union nor the granting authority can be held responsible for them.

**Conflict of interest disclosure:** We declare no competing interests.

### Competing Interest Statement

The authors have declared no competing interest.

### Author Declarations

The Research Ethics Committee of the Finnish Institute for Health and Welfare approved the study protocol. Informed consent is not required for register-based studies in Finland.

